# A systematic literature review of the National Early Warning Score (NEWS)-2 using Prediction Model Risk of Bias Assessment Tool (PROBAST)

**DOI:** 10.1101/2025.03.03.25322780

**Authors:** Cidalia Eusebio, Alison Leary, David Stuckler

## Abstract

**Background:** Identifying patients who are at risk of deteriorating to prevent avoidable complications is a challenge for health management. To support clinical decision-making, the UK introduced the National Early Warning Score 2 (NEWS-2). However, effectiveness in hospitals is not well understood.

**Objective:** To perform a systematic review of external validation studies of NEWS-2.

**Methods:** We performed a systematic review of NEWS-2 and a critical appraisal using PROBAST. Following PRISMA guidelines we searched Medline, Embase, Global Health and Social Policy and Practice for UK NEWS-2 external validation studies from January 2017 to November 2023. PROBAST was used to assess Risk-of-Bias (ROB), segmenting results by population level and model characteristics.

**Results:** 31 validation studies were included in the final sample, including 19 from the UK, nine from Spain, one from Denmark and two from Sweden. 29 out of the 31 studies included were considered to have a high ROB with one having high risk of applicability. The main sources of bias were analytical weaknesses (n=29), outcome measures (n=8), weak inclusion/exclusion of participants (n=5) and weak predictors (n=2).

**Conclusions:** Most external validation studies of NEWS-2 have a high ROB. Future research is needed to address methodological weaknesses, particularly in predictors measurement, the inclusion of outcomes in the predictor measures, missing data, and inadequate calibration. These findings could help inform future Royal College of Physician’s (RCP) policy reviews of NEWS-2.

**Registration:** Not registered, the protocol was not prepared.

**Guarantor of the work:** Cidalia Eusebio

**Social media abstract:** The high risk of bias in studies evaluating NEWS-2 means we should use caution not to rely on NEWS –2 alone for clinical decision making. #PatientSafety @eusebiocidalia @alisonleary1 @davidstuckler

**Impact:**

- NEWS-2 was introduced in UK hospitals in 2017, but its effectiveness is unclear.
- We performed a critical appraisal of NEWS-2 in hospital and pre-hospital settings.
- Despite having a high ROB in the literature, NEWS-2 is utilised in the NHS.
- Clinicians should be cautious about relying solely on NEWS-2 for making decisions.
- Oxygen scores based on increase oxygen requirements should be reevaluated.
- This review supports reinstating the NEWS-2 policy review by the RCP, which was cancelled in 2023.

**Reporting Method:** We adhered to the relevant EQUATOR guidelines.

**Patient or Public Contribution:** There was no patient or public contribution.

## Background

Following the National Patient Safety agency report in 2007, revealing high rates of avoidable in-hospital mortality, the National Institute for Health and Care Excellence recommended the use of Early Warning Scores (EWS) systems to detect patient deterioration based on vital signs scores.^1^ Although these EWS systems were rapidly adopted across the NHS, the lack of national guidance led to variations in EWS models used by hospitals, hindering effective implementation.

To better standardise EWS in the UK, the Royal College of Physicians (RCP) introduced the National Early Warning Score (NEWS), the country’s first standardised EWS system, in 2012. It was established as useful in predicting unfavourable outcomes such cardiac arrest, intensive care unit (ICU) admission, and mortality ^2,3^ and showed promise in predicting mortality in pre-hospital care and the length of hospital stay.^4,5^ This tool was implemented across the NHS.

Yet despite early positive findings, NEWS had limited reliability for specific population groups such as Chronic Obstructive Pulmonary Disease (COPD), spinal injury, and hypercapnic respiratory failure as a result of overestimation of their risk of deterioration.^6,7^ To overcome these shortcomings, the RCP launched NEWS-2 in 2017, incorporating additional parameters (including level of consciousness scoring and a revised SpO2 scale for patients with hypercapnic respiratory failure) to improve the predictive value of patient deterioration.^8^ These changes were also adopted by the NHS.

There are signs that NEWS-2 may be potentially underperforming. Firstly, recent systematic reviews of EWS, not including NEWS-2, concluded that poor methods in external validation studies could lead to the implementation of inferior EWS systems, with false reassurances about their predictive ability and generalisability.^9^ Secondly, there is limited evidence regardingNEWS-2 for non-hospitalised patients even though the NHS started to roll out of NEWS-2 across non-acute settings in other health and social care settings.

This systematic literature review aims to appraise critically, using the Prediction model study Risk of Bias Assessment tool (PROBAST), the validation of external prognostic NEWS-2 studies. Following analysis, we discuss the results and recommendations to support future NEWS-2 review by the RCP.

## Methods

We conducted a systematic literature review following a multi-step process. Initially, one reviewer was responsible for retrieving the initial pool of studies. Two independent reviewers then screened the studies based on pre-defined inclusion and exclusion criteria. After the initial screening all three reviewers engaged in discussions to resolve any discrepancies and reach a consensus. No external data confirmation from study investigators was required.

We used both Preferred Reporting Items for Systematic Reviews and Meta-Analyses (PRISMA) and the Critical Appraisal and data extraction (CHARMS) to supplement PROBAST. This is the recommended tool to support the evaluation of prediction models, assessing Risk of Bias (ROB) and concerns regarding applicability.^10^ Although still prone to variations in classification, it offers the most comprehensive tools and criteria to evaluate predictive models to date. PRISMA (Figure 1), helped to describe systematic review stages while CHARMS offers specific advice on primary prediction modelling studies.

**Fig 1.**
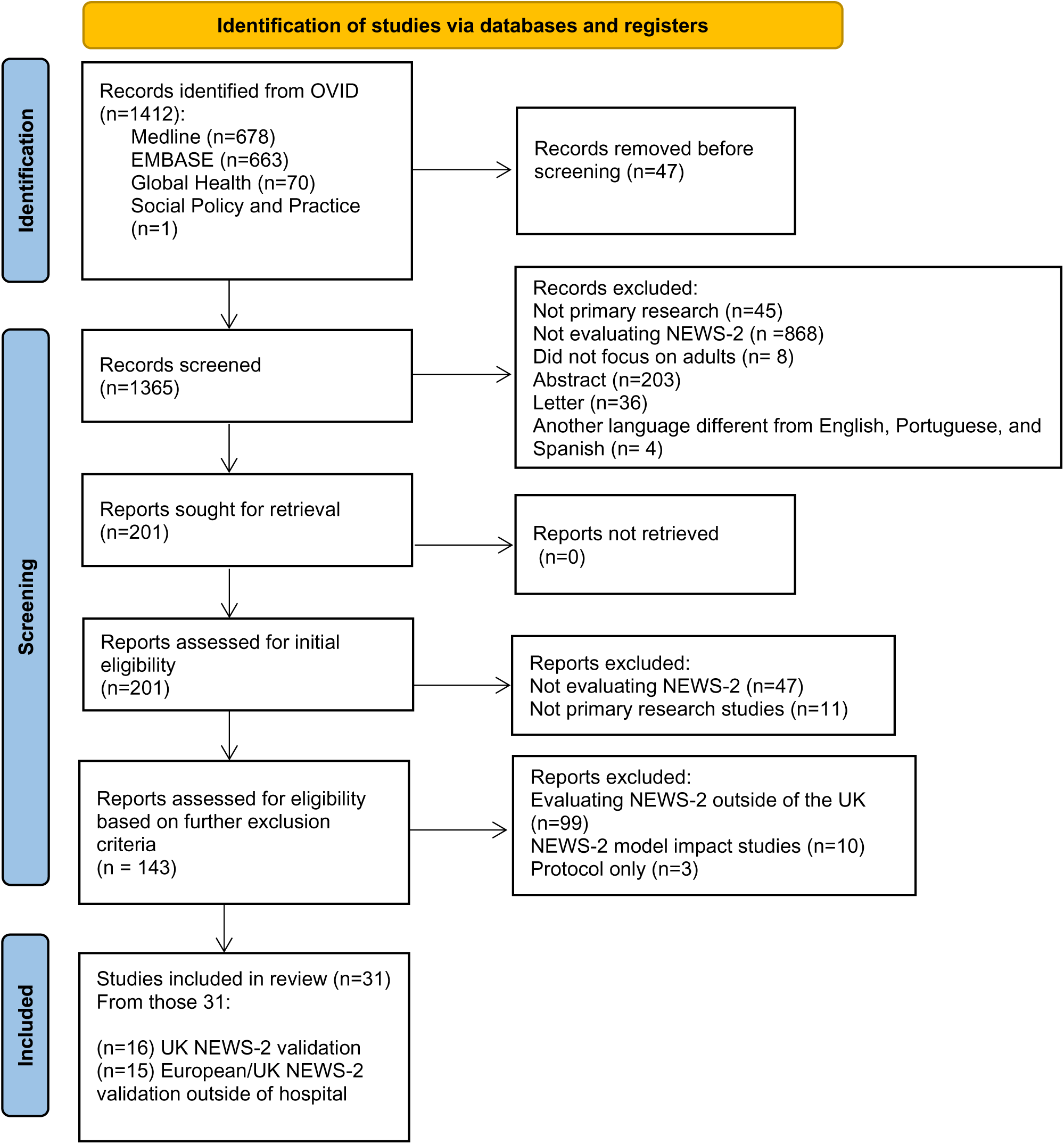
PRISMA Flow Diagram (2007): <u>Diagram of the search process.</u>.

### Databases Searched

We searched the following OVID databases sequentially: MEDLINE, EMBASE, Global Health and Social Policy and Practice for Research for papers on NEWS-2 covering the period from when NEWS-2 was introduced in 2017 through to November 2023.

Our search was supported by a librarian and the following search terms were employed: (National Early Warning Score or NEWS2 or NEWS 2 or National Early Warning Score 2).mp. [mp=title, abstract, original title, name of substance word, subject heading word, floating sub-heading word, keyword heading word, organism supplementary concept word, 15 protocol supplementary concept word, rare disease supplementary concept word, unique identifier, synonyms] limit 1 to yr="2017 -Current"

### Eligibility criteria

Following the guidance of the CHARMS checklist, we established the inclusion and exclusion criteria for selecting studies in this review.^11^ The population studied was adults in the UK in any setting. In addition, due to the lack of studies in non-hospitalised adults in the UK, we included studies in non-hospitalised adults elsewhere between January 2017 and November 2023. NEWS-2 was a index with no comparator used. Primary outcomes were our focus, with no restrictions to capture the full range of ways NEWS-2 was employed. In addition, by focussing on non-hospitalised populations, we anticipate different outcomes from those observed in the internal validation study.

### Study Selection and data extraction

In the initial search across OVID databases, a total of 1412 records were identified. After removing duplicates and screening titles and abstracts, 1365 records remained. We excluded records that were not primary research, did not evaluate NEWS-2, did not focus on adults, were abstracts or letters, or were in languages other than English, Portuguese, or Spanish, due to our proficiency. This led to the exclusion of 1222 articles, leaving 201 papers for further evaluation. An addition screening and exclusion were made to ensure that the papers evaluated NEWS-2 specifically, resulting in 31 articles that met the eligibility criteria.

Once the final studies were extracted, initially, we classified the type of prediction model evaluation as external, internal or both as validation and development need to be assessed independently in the following step.^10^

### Assessment of Bias

Following classification, we assessed the ROB and applicability for each study for domains recommended by the PROBAST tool: Participants, Predictors, Outcome and Analysis. ^10^ The bias classification of the studies is dependent on their low, high, or unclear risk in each domain. The analysis followed the recommended checklist for each domain. This can be viewed in Appendix 1.

## Results

We reviewed 31 articles externally validating NEWS-2, of which four include a model update (including new predictors into NEWS-2). Sixteen of these studies focused on the hospitalised population in the UK, of which nine focused on Covid patients, one on COPD patients, and six on emergency admissions (Figure 2).

**Fig 2.**
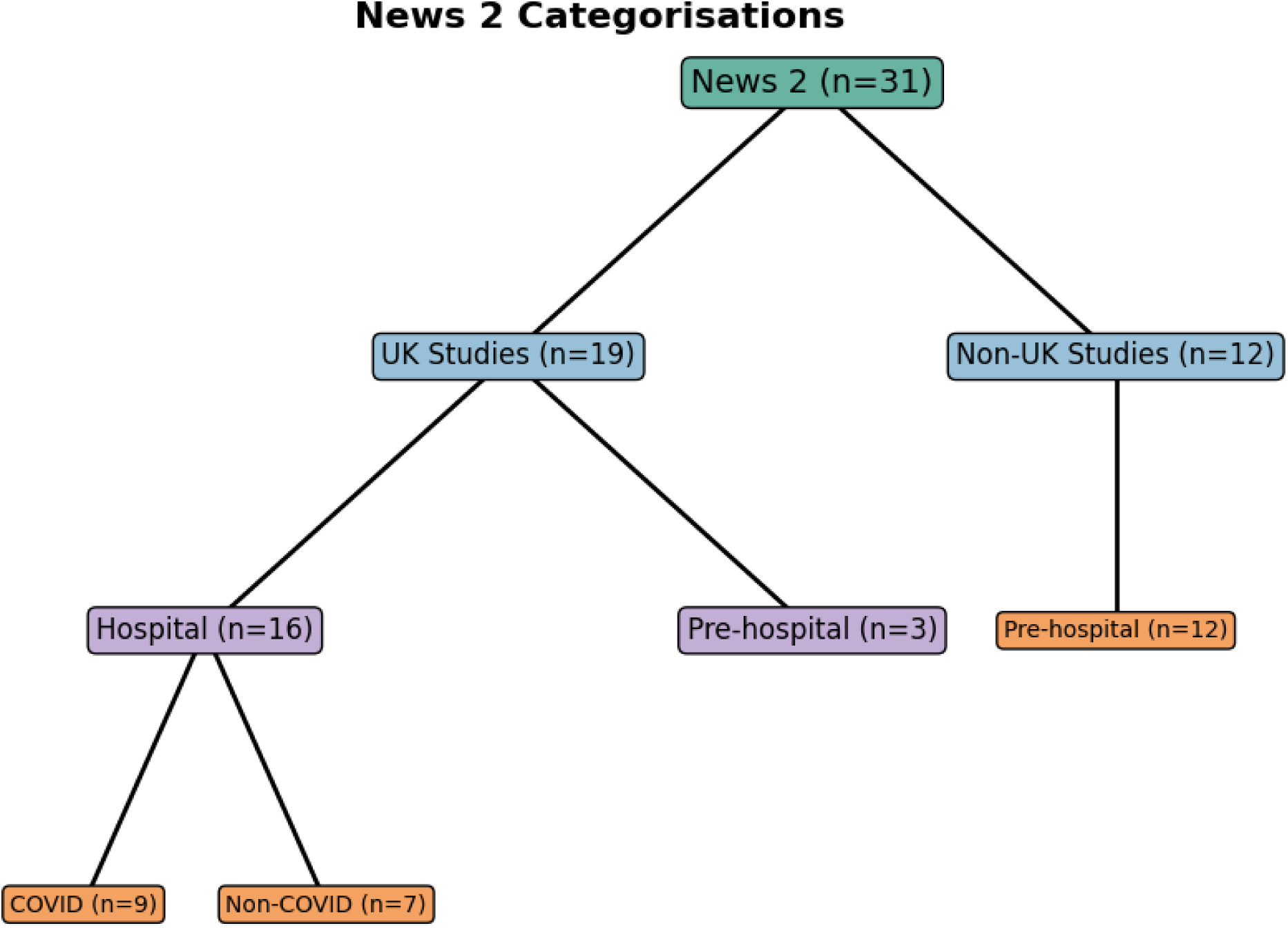
NEWS 2 Categorisations.

Fifteen studies were in prehospital populations. Twelve articles were from outside the UK, including nine from Spain, one from Denmark and two from Sweden. Although most of these studies, (n=24) focused on benchmarking NEWS-2 against other models, this analysis in this study will focus on the validation of NEWS-2. Table 1 provides the study design characteristics and ROB of each study with one issue found on the study’s applicability.

**Table 1.**
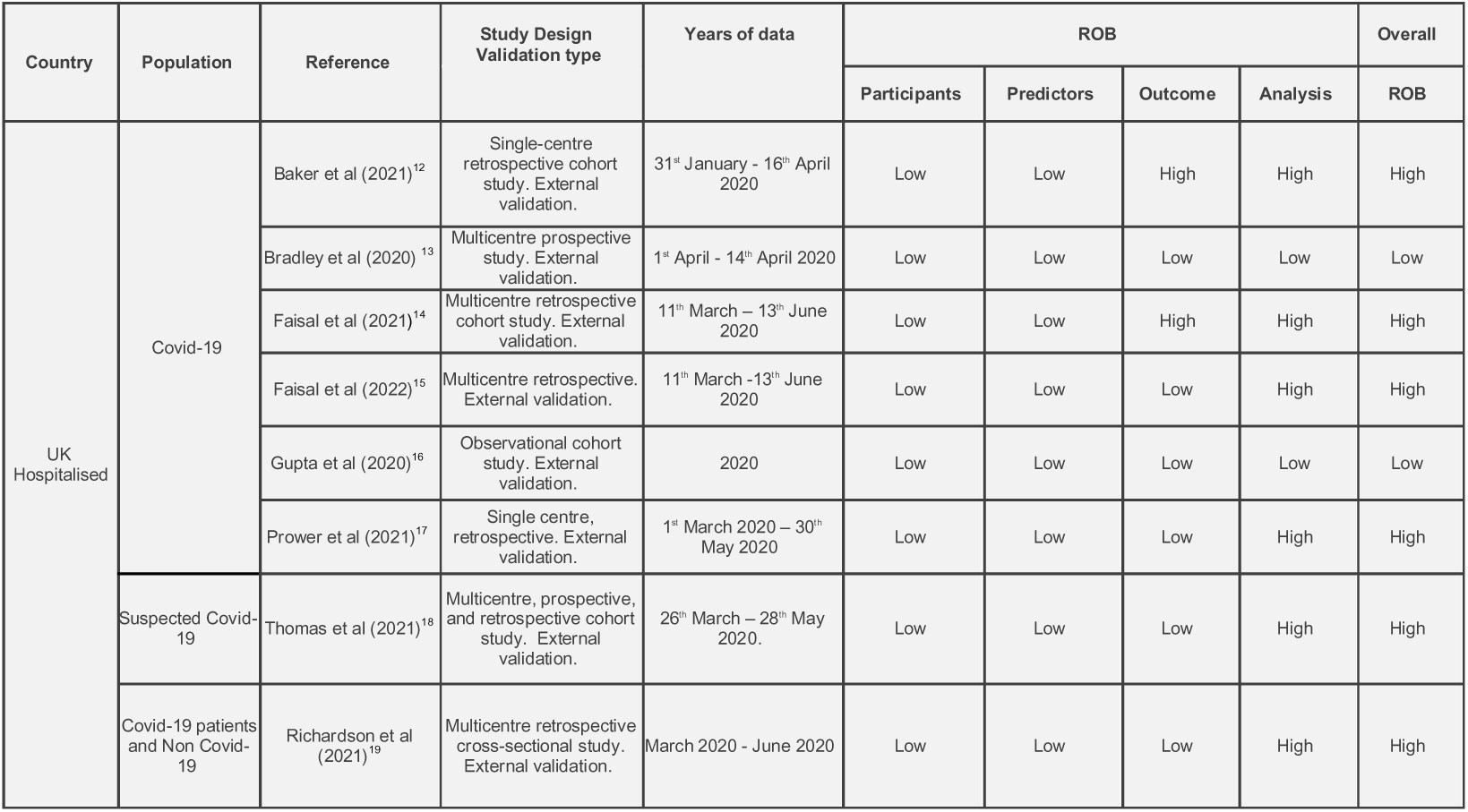
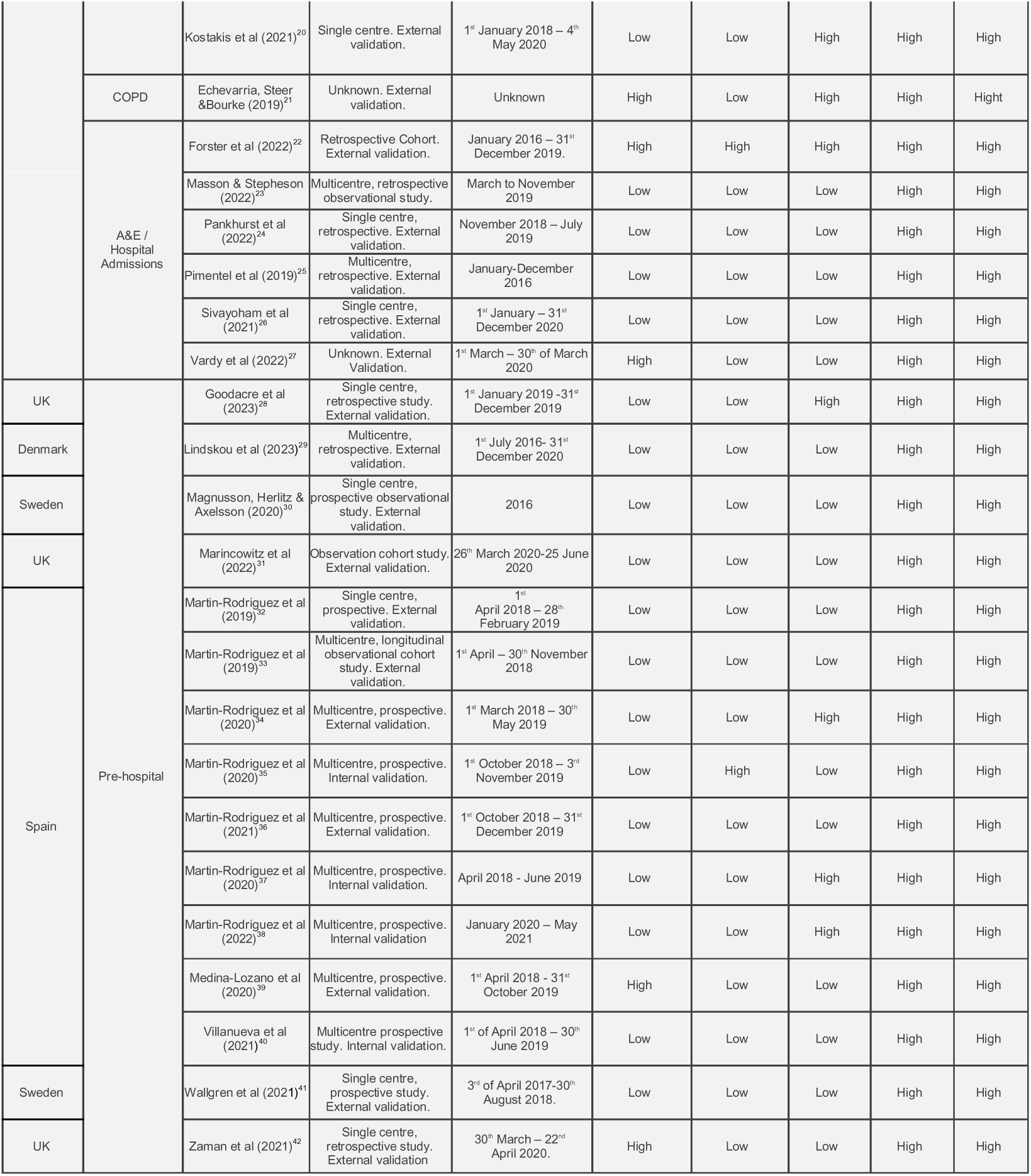
Study design characteristics and ROB of NEWS-2 articles.

### Participants

The majority of studies (n=26) had a low ROB in participant characteristics, consistent with limitations pointed out by the Transparent Reporting of a multivariable prediction model for Individual Prognosis or Diagnosis (TRIPOD). Although the studies used recommended designs, they lacked detailed descriptions or justification for the number of centres, type of hospitals, demographics, and inclusion/exclusion criteria. Several studies (n=9) claiming to validate NEWS-2 in pre-hospital patients lacked representativeness, resulting in the model’s generalizability being challenging.

Both NEWS-2 Covid-19 (n=5) and non-Covid studies (n=7) were mainly retrospective using electronic health records (EHR). In comparison with NEWS-2 studies in a prehospital setting which were mostly prospective cohort studies (n=10). Most Covid-19, non-Covid and prehospital studies offered data at a multicentre level. Areas covered were North-west London, Newcastle upon Tyne (COVID-19), and Oxford (Non-Covid). Those evaluating prehospital NEWS-2 were done mainly central and North of Spain (n=7), Denmark (n=1) and in Sweden (n=2). Studies varied in their data durations and sample sizes.

Five studies had a high ROB among the participants mainly due to inclusion and exclusion criteria and the type of study selected. One study included child data despite focusing on adults, while other studies had limited information regarding the hospitals included, the type of study, and inclusion and exclusion criteria.^18,21,27,39,42^

We observed that studies across these populations, often excluded specific populations like pregnant women, those with mental health issues. Others included only elderly populations or assume that all of the population did not have hypercapnic respiratory failure.^42^ In prehospital studies, some have accounted for situations that would impact the results estimates and excluded those in palliative care, cardiac and respiratory arrest during pre-hospital care and death.^32,39^

### Predictor measures

Two non-Covid-19 studies presented high ROB on the predictor measures due to several factors. One study did not include confusion as a predictor, even though it is a predictor measure of NEWS-2, and itself can suggest deterioration.^25^ Other studies included data prior NEWS-2 availability.^22^ Some Covid-19 studies did not clarify if predictors were measured individually or by NEWS-2 thresholds.^14,23,24^ The majority of studies in the prehospital settings explained and measured confusion following 2017 RCP recommendations. Clinical training and the use of the same equipment for vital signs measurement helped reduce differential misclassification risk.^8^

Only one study was found to have high risk of bias in the applicability as Prehospital NT-proBNP is unlikely to be measured as standard in most prehospital settings.^38^

### Outcome measures

The Covid-19 studies present primary outcomes’ heterogeneity – which differ from primary validated development study of NEWS-2. The most measured outcome was mortality at 30 days (n=12) and mortality at 48 hours (n=10). There are several studies presenting other types of primary outcomes which vary in their description and assessment. Examples included outcomes such as serious event occurrence, intensive care unit admission, deterioration, respiratory or cardiac support and sepsis.^12,16,38^

Relative to ROB, we found 8 studies with high outcomes risk. These ROB were due to, firstly studies choosing different measures of outcomes than the primary study of NEWS-2. Secondly, it is unclear whether the assessment of outcomes was blinded to the predictors.^12,17,19–21^ Finally, we seen the predictors forming part of the definition assessment of the outcome, particularly for the studies assessing sepsis. One study opted for serious event occurrence as an outcome which can create concerns towards the analysis of ROB relative to NEWS-2.^39^

### Analysis

Twenty-nine out of thirty-one studies present high ROB mainly due to failing to include all the enrolled participants in the analysis, and to evaluate calibration.^30,36,41^ The only study assessing calibration, used the Hosmer-Lemeshow goodness-of-fit test and visual assessment of calibration plots.^21^ Other studies presented issues regarding the handling of missing data, presenting classification measures, and not accounting for data complexities.^20,26,28,29,31^

All NEWS-2 Covid-19 studies assessed area under the receiving operating curve (AUROC) with values ranging from (0.65 to 0.89), depending on the outcomes. Regarding NEWS-2 non-Covid-19 studies, two studies, evaluated COPD and type II respiratory failure with AUROC values ranging from (0.70 to 0.81) respectively.^21,25^ For patients admitted to emergency care, AUROC varied with values ranging from (0.86 to 0.96), depending on the outcomes, being mortality at 48 hrs presenting the highest AUROC.

NEWS-2 studies on pre-hospital populations, assessed model discrimination, with AUROC values ranging from (0.52 to 0.91), depending on outcomes. All studies presented classification measures. Besides those limitations, eleven studies had less than 100 participants with the outcome, affecting the model’s predictive performance.^14,15,32,32,34,35,37–40,42^

### Overall PROBAST ROB

Twenty-nine (94%) of 31 studies had high ROB, with the majority presenting analysis limitations(Figure 3). This was followed by high ROB in outcomes, eight articles (26%), participants (16%) with five articles and two studies due to predictors. High ROB in outcomes was mainly due to subjective outcomes. As demonstrated in figure 4, there is significant heterogeneity between outcomes evaluated by studies. This has implications for the comparison of results, especially in Covid-19 populations, with twelve different primary outcomes evaluated.^43^ Pre-hospital studies evaluate mortality at different timeframes, but mainly at 48 hours, which facilitates the comparison of these study results. One study was found to have a high risk to applicability due to the employment of a measurement of NT-proBNP which is unlikely to be measured as standard in community settings.^38^

**Fig 3.**
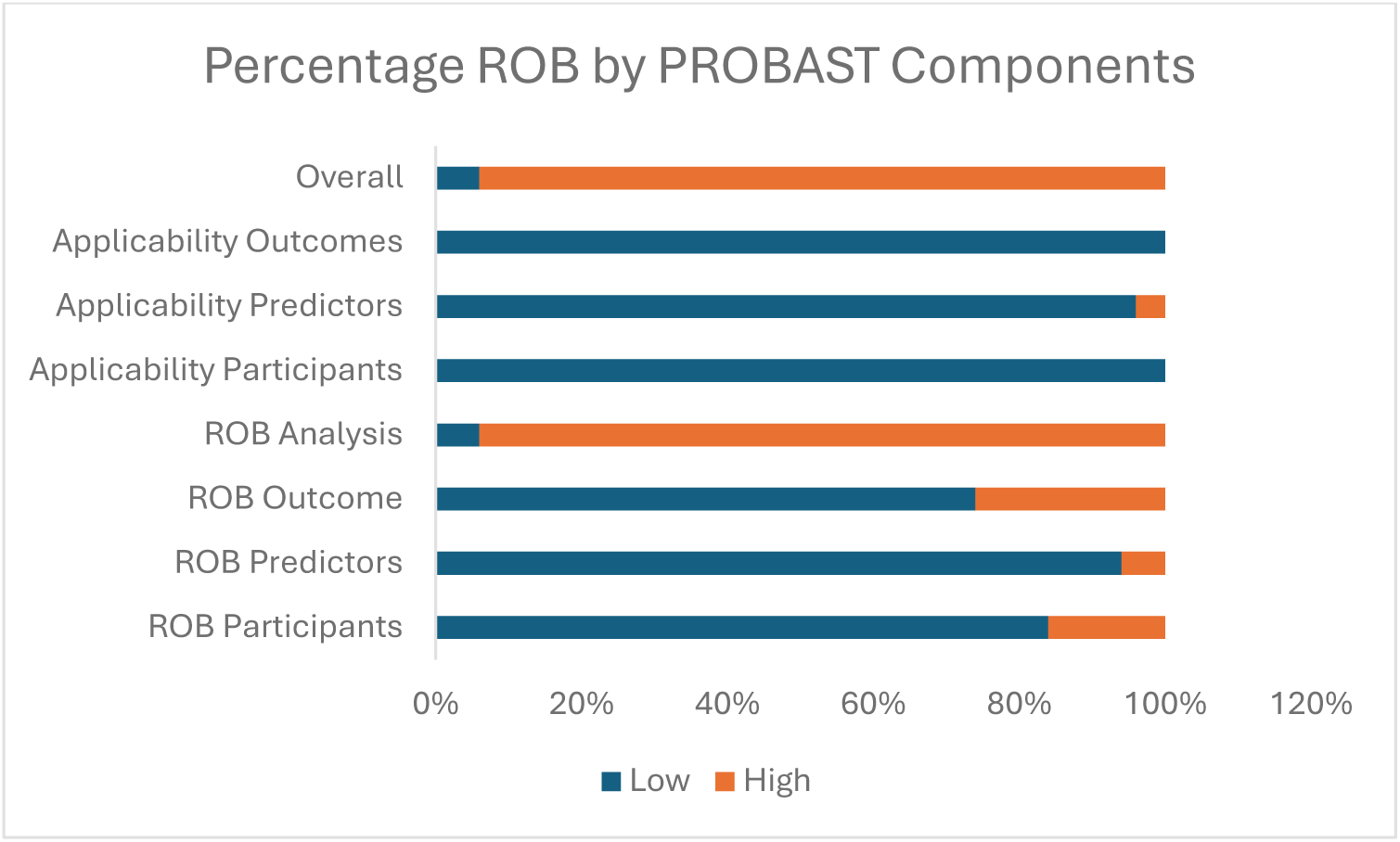
Percentage ROB by PROBAST components.

**Fig 4.**
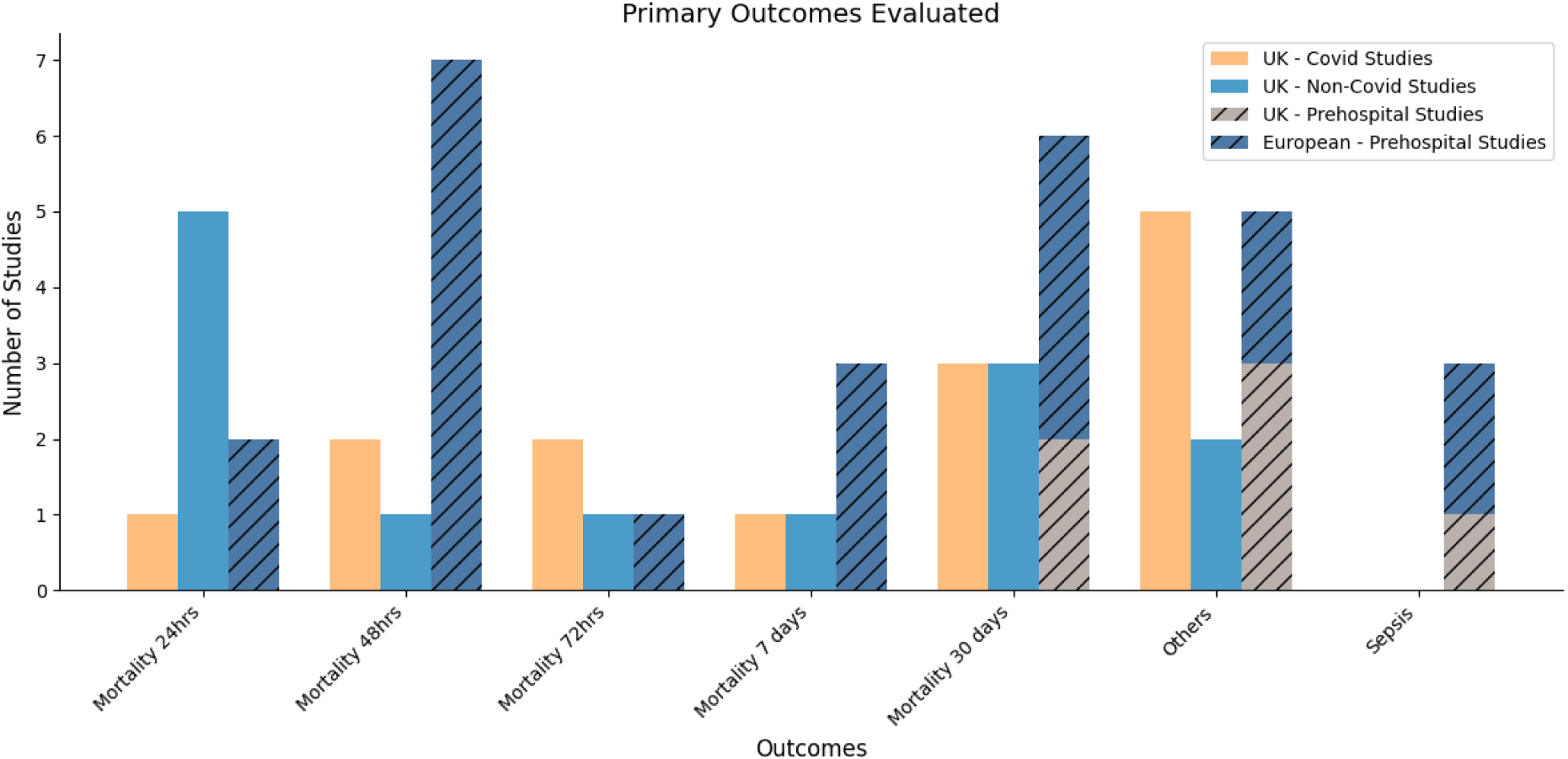
Primary outcomes evaluated.

## Discussion

The review of 31 studies on NEWS-2 validation found numerous methodological issues: 29 were considered to have a high ROB, mostly due to analytical flaws and inadequate outcome measures. The screening process involved multiple stages due to the absence of some of the four key study elements in titles and abstracts recommended by TRIPOD guidelines - the type of modelling study, the clinical context, the target populations, and the outcomes being predicted.^11^

### Participants

Study design, a lack of representativeness, the use of convenience sampling and the impact of exclusion criteria on generalizability are four of the limitations contributing tor the ROB. This aligns with similar findings in the literature.^9,43^ Implementing recommended study designs and considering diverse patient populations could improve the reliability and generalizability of NEWS-2. Often pre-hospital studies excluded pregnant patients and; patients with mental health issues. In some cases they used teams closer to hospital as a convenience sample, which impacts on the representativeness of populations.

Excluding specific populations could also lead to overestimation of outcome risk or restrict applicability. For example, in Covid-19 studies, patients on high flow oxygen were excluded, despite frequent clinical use of this.^17^ This affects model generalizability and presents challenges for clinicians when treating unrepresented populations. Encouraging more comprehensive study designs and transparent reporting of participants can enhance both policy, research, and clinical effectiveness.

### Predictors

Although most studies were considered low ROB, limitations such as incomplete descriptions, unclear validation of predictors and lack of consideration for clinical decision making were found. Vital signs and level of consciousness measurement varied between studies. Some controlled for potential misclassification, while others didn’t, due to the use of retrospective data. This could overestimate NEWS-2 performance in routine clinical practice.^44^

Most of these studies were unclear on how NEWS-2 predictors were validated and which threshold levels were used. In some studies, it is unclear if they evaluate predictors as univariable, categories or in different categories. In addition, it is uncertain whether some studies evaluated the same thresholds levels as the primary study. This absence of information might influence the over or underestimation of results and generate different conclusions.^45^ For example, employing a binary scoring for supplemental oxygen fails to differentiate an increase in oxygen rates, particularly as an increased oxygen requirement should prompt a clinical review.^12,25,43^ Clinicians should be cautious when interpreting and applying predictor models that lack comprehensive validation. To minimize the risk, it is recommended that predictors are continuously monitored to assess their relationship with the outcome.^44^ Furthermore, NEWS-2 thresholds were chosen by clinical consensus rather than evidence, which may omit crucial information.^46^

### Outcomes

Eight studies have presented high ROB in the outcomes, mainly associated with evaluating primary non-mortality outcomes or lack of justification, blinding outcome assessment and limited discussion on confounding and causality. Similar findings have been reported in a previous literature review.^47^ Often outcome choice differed from the original primary development NEWS study. Some studies opted for a varied non-mortality outcome such as, deterioration, sepsis, or septic shock. This presents a deviation of the primary outcome of NEWS-2 but also may result in bias. The utilisation of scores from the Sepsis 3 tool as predictors can potentially lead to overfitting and a reduction in the generalizability of the data.

Outcome is ideally assessed while blinded regarding information about the predictors, but most studies do not report if this is the case.^10,43^ This presents challenges, especially with non-mortality outcomes. Firstly, it’s unethical to omit vital sign measurements, as they are crucial in decision-making. Secondly, NEWS-2 is already integrated into most NHS hospitals’ decision-making processes. Lastly, it can also lead to confounding as having an suitable intervention/prompt clinical care may prevent the undesired outcome from occurring, labelling many patients at risk unnecessarily.^48^

Recent research introduces a ‘predictimand’ framework aiming to account for risk relative to treatment after baseline, as TRIPOD guidance offers limited discussion on confounding, effect modification, and causality.^49^ Ideally, we should understand the independent contribution (e.g. comorbidities) to outcome prediction as factors, as may have an greater impact on the outcome rather than the intervention itself.^44^ The use of electronic medical records might help address some of these limitations in the future.

Transparency and justification of the selection of outcomes, attempting to blind outcome assessment when feasible and the adherence to validated guidelines can help mitigate bias and improve the reliability of study findings. From a clinician’s perspective, understanding these limitations can help clinicians to critically evaluate the applicability of the study findings to their practice.

### Analysis

Analysis proved the most problematic area, with 29 studies demonstrating high ROB. Common factors included lack of calibration measures, incomplete participant enrolment in the analysis, missing data, and lack of information about competing risks affecting outcomes.

While calibration assessment is widely recommended, it is often the component least evaluated. It is important to improve this evaluation, as poorly calibrated algorithms can mislead and potentially harm clinical decision-making.^50^ Furthermore, studies measuring calibration studies reported overfitting results, which is concerning, as NEWS-2 is widely implemented across the NHS.

Including only complete data in studies is inefficient. Firstly, it reduces the sample size. Secondly, the results may be biased, increasing the samples’ risk of not being representative of the population. Finally, it may lead to different bias estimates of the predictor/outcome associations on the model predictor performance compared to those what would be obtained if the whole set of data was to be analysed.^50^

Some studies did not present a reasonable number of outcomes per participant. The number of outcomes required is an active area of research. While the TRIPOD guidance suggests more than 250 outcome events in external validation studies, a recent study, suggests using a simulation-based approach to calculate sample size.^45,50^

### Implications for future RCP NEWS-2 review

This NEWS-2 literature review offers insights to improve policy, research and clinical practice. From a policy perspective, although the NHS intends to implement NEWS-2 across health and social care, evidence remains scarce for prehospital care in the UK. In addition, any policy should encourage the application of standard guidelines, comprehensive study designs and transparent reporting of participants.

To enhance future research, it is advisable to follow the recommendations by Gerry and colleagues and those outlined in TRIPOD.^9,45^ Leveraging the wealth of data from electronic health records (EHR) implemented across the NHS can foster more effective, complex models, contributing to our understanding of NEWS-2 in other contexts as well as non-linear complex relationships between outcomes, economic benefits and other unknow relationships.

Triggers selection should be evidence based. The binary scoring for oxygen delivery should be reevaluated and continuous prediction evaluation is recommended. Training on NEWS-2 should address its high ROB flaws, enabling healthcare professionals to better comprehend its limitations.

## Limitations

This review has some limitations. Our database might be bias towards North American and European journals and studies in languages including English, Portuguese and Spanish. Based largely on UK data, this review does not compare other EWS or extend to other countries. The heterogeneity of studies also limits general applicability. Inclusion of early pandemic Covid-19 data and external studies primarily from a specific area in Spain may present biases. In addition, while the PROBAST tool provides specific criteria for assessing the risk of bias, there remains the potential for subjective interpretation by researchers.

## Strengths

This literature review has several strengths. Firstly, it provides specific considerations tailored to researchers, policymakers, and clinicians, to effectively navigate methodological limitations. Secondly, is the only literature review on NEWS-2 critically appraising the ROB beyond Covid-19 patients. Is also the first literature review looking at the evidence of NEWS-2 in non-hospital settings. In addition, implications for research, policy and management are briefly discussed helping policymakers with NEWS-2 future revisions. Our study corroborated the observations of previous literature reviews who also evaluated alternative EWS, including NEWS, but not NEWS-2.^9^

## Conclusions

The literature review has critically assessed the ROB and applicability of NEWS-2 in the UK and for non-UK pre-hospitalised patients using the PROBAST tool. Analysis found evidence of many methodological and reporting shortcomings, that might impact the overall findings of the studies and have implications for practice and patient safety.

29 (94%) out of 31 studies were considered high ROB, all presenting analysis limitations. This followed by outcomes with, eight articles (25%), participants (16%) with five articles and two studies due to predictors. The high ROB in outcomes was mainly related to having a subjective outcome where this is determined with the predictor’s knowledge. One study had high risk of applicability.

This study is the first literature review on NEWS-2. The findings of the review align with previous EWS systematic reviews, warning that poor methods in external validation studies could lead to the implementation of inferior EWS systems, with false reassurances about their predictive ability and generalisability.^9,43,51,52^

A key concern which should suggest a review of NEWS-2 is the underestimation of patient deterioration due to oxygen binary score scales and respiratory predictors. Improved methodology guidance from TRIPOD and greater understanding of NEWS-2 flaws amongst health professionals are recommended until more accurate prediction models emerge from EHR.

Clinicians should recognise that competency-based approaches should always be balanced with professional judgement. Whist NEWS-2 is a decision-making facilitating tool embedded in a competency-based workforce; it should not undermine clinical judgement. It can help with decision-making, not because of the model effectiveness, rather because it streamlines an operational process to trigger a health system response. This Tayloristic perspective embedded in the modern healthcare, oversimplifies work as series of uncontextualized independent tasks, overlooking the complexity and the critical thinking that is an essential part of practice as a healthcare professional.^53^

## Supporting information

Supplemental PROBAST guidance

## Data Availability

All data produced in the present work are contained in the manuscript

## Acknowledgments

We thank Assistant Professor Dr Thomas Cowling and Professor Jose Neves for providing comments on the manuscript.

## Ethical approval

This is a review of publicly available studies and as such was not submitted for ethical review.

## Consent to participate

Not applicable

## Data availability statement

This is a review. Data is in the public domain.

