## Supplemental PROBAST guidance for "A systematic literature review of the National Early Warning Score (NEWS)-2 using Prediction Model Risk of Bias Assessment Tool (PROBAST)"

### Appendix 5: PROBAST Checklist tool by Moons et al. (2019)

1. PROBAST: A Tool to Assess the Risk of Bias and Applicability of Prediction Model Studies
2. PROBAST: A Tool to Assess Risk of Bias and Applicability of Prediction Model Studies: Explanation and Elaboration

**What does PROBAST assess?**
PROBAST assesses both the *risk of bias* and *concerns regarding applicability* of a study that evaluates (develops, validates or updates) a multivariable diagnostic or prognostic prediction model. It is designed to assess primary studies included in a systematic review.

*Bias* occurs if systematic flaws or limitations in the design, conduct or analysis of a primary study distort the results. For the purpose of prediction modelling studies, we have defined *risk of bias* to occur when shortcomings in the study design, conduct or analysis led to systematically distorted estimates of a model's predictive performance or to an inadequate model to address the research question. Model predictive performance is typically evaluated using calibration, discrimination and sometimes classification measures, and these are likely inaccurately estimated in studies with high risk of bias. *Applicability* refers to the extent to which the prediction model from the primary study matches your systematic review question, for example in terms of the participants, predictors or outcome of interest.

A primary study may include the development and/or validation or update of more than one prediction model. A PROBAST assessment should be completed for each distinct model that is developed, validated or updated (extended) for making individualised predictions. Where a publication assesses multiple prediction models, only complete a PROBAST assessment for those models that meet the inclusion criteria for your systematic review. Please note that subsequent use of the term "model" includes derivatives of models, such as simplified risk scores, nomograms, or recalibrations of models.

PROBAST is not designed for all multivariable diagnostic or prognostic studies. For example, studies using multivariable models to identify predictors associated with an outcome but not attempting to develop a model for making individualised predictions are not covered by PROBAST.

PROBAST includes four steps.

| Step | Task | When to complete |
|---|---|---|
| **1** | Specify your systematic review question(s) | Once per systematic review |
| **2** | Classify the type of prediction model evaluation | Once for each model of interest in each publication being assessed, for each relevant outcome |
| **3** | Assess risk of bias and applicability | Once for each development and validation of each distinct prediction model in a publication |
| **4** | Overall judgment | Once for each development and validation of each distinct prediction model in a publication |

If this is your first time using PROBAST, we strongly recommend reading the detailed explanation and elaboration (E&E, see link above) paper and to check the examples on www.probast.org

**Step 1: Specify your systematic review question**

State your systematic review question to facilitate the assessment of the applicability of the evaluated models to your question. *The following table should be completed once per systematic review.*

| **Criteria** | **Specify your systematic review question** |
|---|---|
| *Intended use of model:* | |
| ***Participants** including selection criteria and setting:* | |
| ***Predictors** (used in prediction modelling), including types of predictors (e.g. history, clinical examination, biochemical markers, imaging tests), time of measurement, specific measurement issues (e.g., any requirements/ prohibitions for specialized equipment):* | |
| *Outcome to be predicted:* | |

**Step 2: Classify the type of prediction model evaluation**

Use the following table to classify the evaluation as model development, model validation or model update, or combination. Different signalling questions apply for different types of prediction model evaluation. If the evaluation does not fit one of these classifications, then PROBAST should not be used.

| **Classify the evaluation based on its aim** | | | |
|---|---|---|---|
| **Type of prediction study** | **PROBAST boxes to complete** | **Tick as appropriate** | **Definition for type of prediction model study** |
| Development only | Development | | Prediction model development without external validation. These studies may include internal validation methods, such as bootstrapping and cross-validation techniques. |
| Development and validation | Development and validation | | Prediction model development combined with external validation in other participants in the same article. |
| Validation only | Validation | | External validation of existing (previously developed) model in other participants. |

*This table should be completed once for each publication being assessed and for each relevant outcome in your review.*

| |
|---|
| **Publication reference** |
| **Models of interest** |
| **Outcome of interest** |

**Step 3: Assess risk of bias and applicability**

PROBAST is structured as four key domains. Each domain is judged for risk of bias (low, high or unclear) and includes signalling questions to help make judgements. Signalling questions are rated as yes (Y), probably yes (PY), probably no (PN), no (N) or no information (NI). All signalling questions are phrased so that "yes" indicates absence of bias. Any signalling question rated as "no" or "probably no" flags the potential for bias; you will need to use your judgement to determine whether the domain should be rated as "high", "low" or "unclear" risk of bias. The guidance document contains further instructions and examples on rating signalling questions and risk of bias for each domain.
The first three domains are also rated for concerns regarding applicability (low/ high/ unclear) to your review question defined above.
*Complete all domains separately for each evaluation of a distinct model. Shaded boxes indicate where signalling questions do not apply and should not be answered.*

| | | | |
|---|---|---|---|
| **DOMAIN 1: Participants** | | | |
| **A. Risk of Bias** | | | |
| *Describe the sources of data and criteria for participant selection:* | | | |
| | | Dev | Val |
| 1.1 Were appropriate data sources used, e.g. cohort, RCT or nested case-control study data? | | | |
| 1.2 Were all inclusions and exclusions of participants appropriate? | | | |
| **Risk of bias introduced by selection of participants** | **RISK:** *(low/ high/ unclear)* | | |
| *Rationale of bias rating:* | | | |
| **B. Applicability** | | | |
| *Describe included participants, setting and dates:* | | | |
| **Concern that the included participants and setting do not match the review question** | **CONCERN:** *(low/ high/ unclear)* | | |
| *Rationale of applicability rating:* | | | |

| DOMAIN 2: Predictors | | | |
|---|---|---|---|
| **A. Risk of Bias** | | | |
| *List and describe predictors included in the final model, e.g. definition and timing of assessment:* | | | |
| | | Dev | Val |
| 2.1 Were predictors defined and assessed in a similar way for all participants? | | | |
| 2.2 Were predictor assessments made without knowledge of outcome data? | | | |
| 2.3 Are all predictors available at the time the model is intended to be used? | | | |
| **Risk of bias introduced by predictors or their assessment** | **RISK:** *(low/ high/ unclear)* | | |
| *Rationale of bias rating:* | | | |
| **B. Applicability** | | | |
| Concern that the definition, assessment or timing of predictors in the model do not match the review question | **CONCERN:** *(low/ high/ unclear)* | | |
| *Rationale of applicability rating:* | | | |

| DOMAIN 3: Outcome | | | |
|---|---|---|---|
| **A. Risk of Bias** | | | |
| *Describe the outcome, how it was defined and determined, and the time interval between predictor assessment and outcome determination:* | | | |
| | | Dev | Val |
| 3.1 Was the outcome determined appropriately? | | | |
| 3.2 Was a pre-specified or standard outcome definition used? | | | |
| 3.3 Were predictors excluded from the outcome definition? | | | |
| 3.4 Was the outcome defined and determined in a similar way for all participants? | | | |
| 3.5 Was the outcome determined without knowledge of predictor information? | | | |
| 3.6 Was the time interval between predictor assessment and outcome determination appropriate? | | | |
| **Risk of bias introduced by the outcome or its determination** | **RISK:** *(low/ high/ unclear)* | | |
| *Rationale of bias rating:* | | | |

| **B. Applicability** | |
|---|---|
| *At what time point was the outcome determined:*<br>*If a composite outcome was used, describe the relative frequency/distribution of each contributing outcome:* | |
| **Concern that the outcome, its definition, timing or determination do not match the review question** | **CONCERN:***(low/ high/ unclear)* |
| *Rationale of applicability rating:* | |

| **DOMAIN 4: Analysis** | | |
|---|---|---|
| **Risk of Bias** | | |
| *Describe numbers of participants, number of candidate predictors, outcome events and events per candidate predictor:* | | |
| *Describe how the model was developed (for example in regard to modelling technique (e.g. survival or logistic modelling), predictor selection, and risk group definition):* | | |
| *Describe whether and how the model was validated, either internally (e.g. bootstrapping, cross validation, random split sample) or externally (e.g. temporal validation, geographical validation, different setting, different type of participants):* | | |
| *Describe the performance measures of the model, e.g. (re)calibration, discrimination, (re)classification, net benefit, and whether they were adjusted for optimism:* | | |
| *Describe any participants who were excluded from the analysis:* | | |
| *Describe missing data on predictors and outcomes as well as methods used for missing data:* | | |
| | Dev | Val |
| 4.1 Were there a reasonable number of participants with the outcome? | | |
| 4.2 Were continuous and categorical predictors handled appropriately? | | |
| 4.3 Were all enrolled participants included in the analysis? | | |
| 4.4 Were participants with missing data handled appropriately? | | |
| 4.5 Was selection of predictors based on univariable analysis avoided? | | |
| 4.6 Were complexities in the data (e.g. censoring, competing risks, sampling of controls) accounted for appropriately? | | |
| 4.7 Were relevant model performance measures evaluated appropriately? | | |
| 4.8 Were model overfitting and optimism in model performance accounted for? | | |
| 4.9 Do predictors and their assigned weights in the final model correspond to the results from multivariable analysis? | | |

| **Risk of bias introduced by the analysis** | **RISK:***(low/ high/ unclear)* |
|---|---|
| *Rationale of bias rating:* | |

**Step 4: Overall assessment**

Use the following tables to reach overall judgements about risk of bias and concerns regarding applicability of the prediction model evaluation (development and/or validation) across all assessed domains.
*Complete for each evaluation of a distinct model.*

| **Reaching an overall judgement about risk of bias of the prediction model evaluation** | |
|---|---|
| **Low risk of bias** | If all domains were rated low risk of bias.<br>If a prediction model was developed without any external validation, and it was rated as low risk of bias for all domains, consider downgrading to **high risk of bias**. Such a model can only be considered as low risk of bias, if the development was based on a very large data set and included some form of internal validation. |
| **High risk of bias** | If at least one domain is judged to be at **high risk of bias**. |
| **Unclear risk of bias** | If an unclear risk of bias was noted in at least one domain and it was low risk for all other domains. |

| **Reaching an overall judgement about applicability of the prediction model evaluation** | |
|---|---|
| **Low concerns regarding applicability** | If low concerns regarding applicability for all domains, the prediction model evaluation is judged to have **low concerns regarding applicability**. |
| **High concerns regarding applicability** | If high concerns regarding applicability for at least one domain, the prediction model evaluation is judged to have **high concerns regarding applicability**. |
| **Unclear concerns regarding applicability** | If unclear concerns (but no "high concern") regarding applicability for at least one domain, the prediction model evaluation is judged to have **unclear concerns regarding applicability** overall. |

| **Overall judgement about risk of bias and applicability of the prediction model evaluation** | |
|---|---|
| **Overall judgement of risk of bias** | **RISK:***(low/ high/ unclear)* |
| *Summary of sources of potential bias:* | |
| **Overall judgement of applicability** | **CONCERN:***(low/ high/ unclear)* |
| *Summary of applicability concerns:* | |
